# Mitigating isolation: further comparing the effect of LFD testing for early release from self-isolation for COVID-19 cases

**DOI:** 10.1101/2022.01.25.22269818

**Authors:** Declan Bays, Timothy Whiteley, Hannah Williams, Thomas Finnie, Nick Gent

## Abstract

In a recent paper, we described how lateral flow device (LFD) testing might be used to reduce the amount of excess time individuals spend in isolation following confirmation of a COVID-19 infection. Through the work presented here, we look to expand upon this and explore in more detail the benefit that such an approach might provide. We use our previously described model to study scenarios through the metrics “proportion released still infectious”, “excess time spent in isolation” (time isolated while no longer infectious), and “time spent infectious after early release”. We also look to consider the effect on these metrics by comparing values obtained when a single negative LFD test is required for early release, versus requiring two and three sequential negative LFD tests. Results show that jointly employing self-isolation and LFD testing may deliver sizeable reductions to the proportion of individuals being release while still infection, the average amount of excess time spent in isolation by those no longer a public health threat, and the average amount of time spent infectious by those released early. These effects considered in conjunction could provide a considerable decrease in the public health risk posed by still infectious individuals being released back into the population by actively monitoring their infection status throughout their isolation period. Such an approach could also help lighten the impact incurred on the individual by reducing the amount of time spent in isolation while posing no further public health risk, in addition to alleviating pressures on the economy and in healthcare settings caused by mass isolation in times of high prevalence.

## Introduction

The B.1.1.529 variant of the SARS-CoV-2 virus, since named Omicron, was first reported to the World Health Organisation on 24^th^ November 2021 following identification in South Africa[1]. Omicron sparked initial fears of increased transmissibility due to the large number of mutations, a significant proportion of which are present on the spike protein[2]. As Omicron cases began to be reported internationally, case numbers saw a significant increase. The UK reported its first two confirmed Omicron case on 27^th^ November 2021. Two weeks later, there had been a total of 4,713 confirmed Omicron cases; this subsequently increased to 159,932 confirmed Omicron cases by 27^th^ December 2021[3]. The sharp influx in case numbers raised concerns that currently imposed self-isolation policies may cause a massive labour shortage, in addition to having a considerable economic cost. With healthcare providers and supply chains already under pressure, alternative approaches to self-isolation were sought which might alleviate some of these strains.

In a previous work, we demonstrated how lateral flow device (LFD) testing might be used to monitor the infectious status of isolating individuals[4]. This therefore allows for a tailored self-isolation period where individuals may be released once deemed to no longer be infectious, and thus no longer a public health risk. In this paper, we use the previously described model to look at potential strategies in more detail. We compare the benefits and risks of using a single LFD negative result against the requiring of multiple sequential LFD negative results to permit early release. Through the existing model, we compare scenarios by the metrics “proportion released while infectious “, “average excess hours spent in isolation” and “average time spent infectious following release”. Through this work, we hope to present a stronger case for managing confirmed cases on an individual basis. We do not consider the use of PCR testing in this work due to variable turn-round times and lacking capacity for the purposes envisaged herein.

## Methods

Our model uses a similar Monte Carlo based structure as presented by Bays et al for use in studying border screening policies[4], [5]. The model starts by simulating 500,000 individuals, all being assumed to have become infected with COVID-19 at some point prior, each of which is then randomly assigned an infectious period (given in days), *t*_inf_, sampled from a gamma distribution (shape = 2, scale = 2.1) which has been fitted to real-time data using methods described by Birrell et al[6]. At time *t =* 0, individuals are assumed to have been identified as COVID-19 positive, either through testing or the displaying of symptoms, at which point they enter self-isolation. Time *t =* 0 is also assumed to coincide with the start of each individual’s infectious period. The model then proceeds through the pre-determined self-isolation policy.

Each policy is defined by an initial isolation “burn-in” period, following which individuals may then take regular LFD tests at 24-hour intervals, up until a maximum isolation period of 10 days (240 hours) from the initial onset of infectiousness. In our considered scenarios, regular testing is not permitted to start any earlier than day one (24 hours after admission into isolation); this therefore implies that individuals may not be released earlier day one, day two or day three in scenarios where one, two and three consecutive negatives are required respectively (due to requirement that consecutive tests be taken 24 hours apart) – in the below plots, a grey dashed line is used to represent the inability of individuals to be released any earlier than these times. If individuals remain in isolation after 10 full days, they are then assumed to be released regardless of infection status (as defined in UK policy as of writing[7]). We clarify that days are defined as a fixed 24 hour period, so that in a scenario where individuals must isolate for 5 days before being eligible to take their first LFD test, they must have been isolating for 120 hours – waiting a further 24 hours to undergo a second test if required, etc. This avoids possible variation in isolation times which could emerge were we to use calendar days. A policy may require one, two or three sequential negatives to permit early release (scenario depending). Following recent studies, we use LFD testing positivity as a proxy for whether individuals are currently infectious[8] – a true negative result after infection may then be regarded as indicating “recovery”, where viral load is low enough to no longer be a public health risk. We assume LFD tests have a sensitivity of 80% and a specificity of 100%. This is a conservative choice following research undertaken jointly by UKHSA (then Public Health England) and the University of Oxford[9].

For each LFD test administered on day *t*_i_, if *t*_i_ < *t*_inf_ the individual is regarded as still being infectious. In such a case, a weighted coin toss with an 80% probability of success is performed to determine whether the administered test would provide the individual with a true positive or false negative result – this encapsulates the above described sensitivity rate. However, if *t*_i_ ≥ *t*_inf_, the individual is deemed to have “recovered” and no longer be infectious. This test will then relay a negative result. In the considered scenarios where a single negative result is required for early release, a negative result will lead to the individual exiting self-isolation irrespective of the individual’s true infection status (i.e. recovered or not). In the other considered scenarios, we require 2 or 3 consecutive negative results for individuals to be released early and so past results are tracked appropriately to simulate this. Upon release, the model will then determine the excess time that the individual spend in isolation (i.e. time in isolation while being no longer infectious, given by the difference *t*_*i*_−*t*_inf_) if *t*_*i*_ ≥ *t*_inf_, or the time that the individual will remain infectious following release (given by the difference *t*_inf_−*t*_*i*_) if *t*_*inf*_ ≥ *t*_i_. These individuals then exit the model. And as stated above, those individuals which are still in isolation on day 10 are assumed to be released regardless of infection status. We again calculate either the excess time they spent in isolation or the time they will remain infectious post release. This process is repeated for all simulated individuals, where we total the excess time spent in isolation and the time that will be spent infectious post release. We then obtain the average of both, taking our denominators to be 500,000, to give the expected amount of time that each isolating individual might expect to spend in isolation following recovery, or remain infectious outside of isolation following release. These values then give us a metric by which we can assess the benefit and risk of each scenario. For a final comparison, we also run our model where no testing is in place and individuals are simply released only after a fixed period of isolation.

## Results

To make scenarios comparable, we categorise by “minimum isolation period”, by which we mean the earliest possible time individuals might be released providing predefined conditions are met (i.e. one/two/three negative LFD tests). We believe this to be the most appropriate way of associating scenarios given the added structure that the differing number of required negative tests provides. For example, we associate the scenario which is defined as 5 days of burn-in self-isolation with a single negative test required for early release, with the 4 and 3 days of burn-in self-isolation with two and three negative tests required for early release respectively (as in all these scenarios individuals may be released from day 5). We also reiterate that we define a day of isolation to be a fixed 24-hour period from initial point of isolation; by a 5-day isolation period we mean individuals have spent 120 hours in quarantine and so forth. This avoids possible variation in isolation times which could emerge were we to use calendar days (isolating for 5 calendar days from 01:00 Wednesday, is not the same as isolating for 5 calendars days from 23:00 Wednesday).

For later comparison, we start by using our model to calculate the rates at which we would expect infectious individuals to be released if the management of cases was defined solely as a fixed period of self-isolation (i.e. not undergoing any LFD testing prior to reintroduction):

Additionally, the average amount of time that infectious individuals would be expected to remain infectious following early release and excess time spent in isolation under these blanket approaches:

We then proceed to use our model to determine the expected released rates in scenarios where LFD testing may be used to permit early release should a negative result be returned:

We can further break these results down and look at the days on which infectious individuals are released in each of these scenarios:

Next, we consider the average amount of time that infectious individuals would be expected to remain infectious following early release and excess time spent in isolation under each of these scenarios:

And lastly, the number of tests which would be expected to be used by each person enter self-isolation under all the considered scenarios:

## Discussion

As our results show, in comparison to a blanket approach of mandating all individuals self-isolate for a fixed period, using LFD testing in conjunction with isolation can markedly decrease the proportion on individuals who are released while still infectious. For instance, if we compare the fixed 5-day isolation period approach to the scenario where individuals may be released after 5 days of isolation with one negative LFD result, we see a 46.5% decline in infectious releases. This is further improved by the requirement of additional negative LFD results, giving a 74.0% and 81.4% decrease when two and three sequential negatives are required respectively. Obviously, some decrease is to be expected as we are putting in place additional restraints, but the magnitude of this reduction shows that this provides an attractive and attainable alternative. Of course, as the length of isolation period increases we would expect the overall risk of infectious release to decrease, owing to the fact that we would expect a larger proportion of those isolating to have recovered by the time of initial testing/release. However, evidence of the added benefit of requiring a negative LFD test for release remains; comparing a fixed 8-day isolation period to release on day 8 on condition of one, two or three negative LFD results provides a 37.4%, 50.5% and 52.3% decrease in infectious releases respectively.

A caveat of an early release approach as opposed to a longer term fixed isolation period is that, due to the false negativity rate of LFD tests, individuals may be released while infectious where they might have recovered before the end of a longer term fixed isolation period. This is demonstrated in figure 2, where the shorter the minimum isolation period (i.e. earlier we start allowing LFD tests to be performed) the larger the proportion of infectious individuals released soon after. Nevertheless, we can also see that requiring additional negative tests decreases this effect. Ultimately, this is a trade-off between risk of infectious release and number of LFD negatives required for early release, which would have to be adjusted to suit risk appetite.

**Figure 1:**
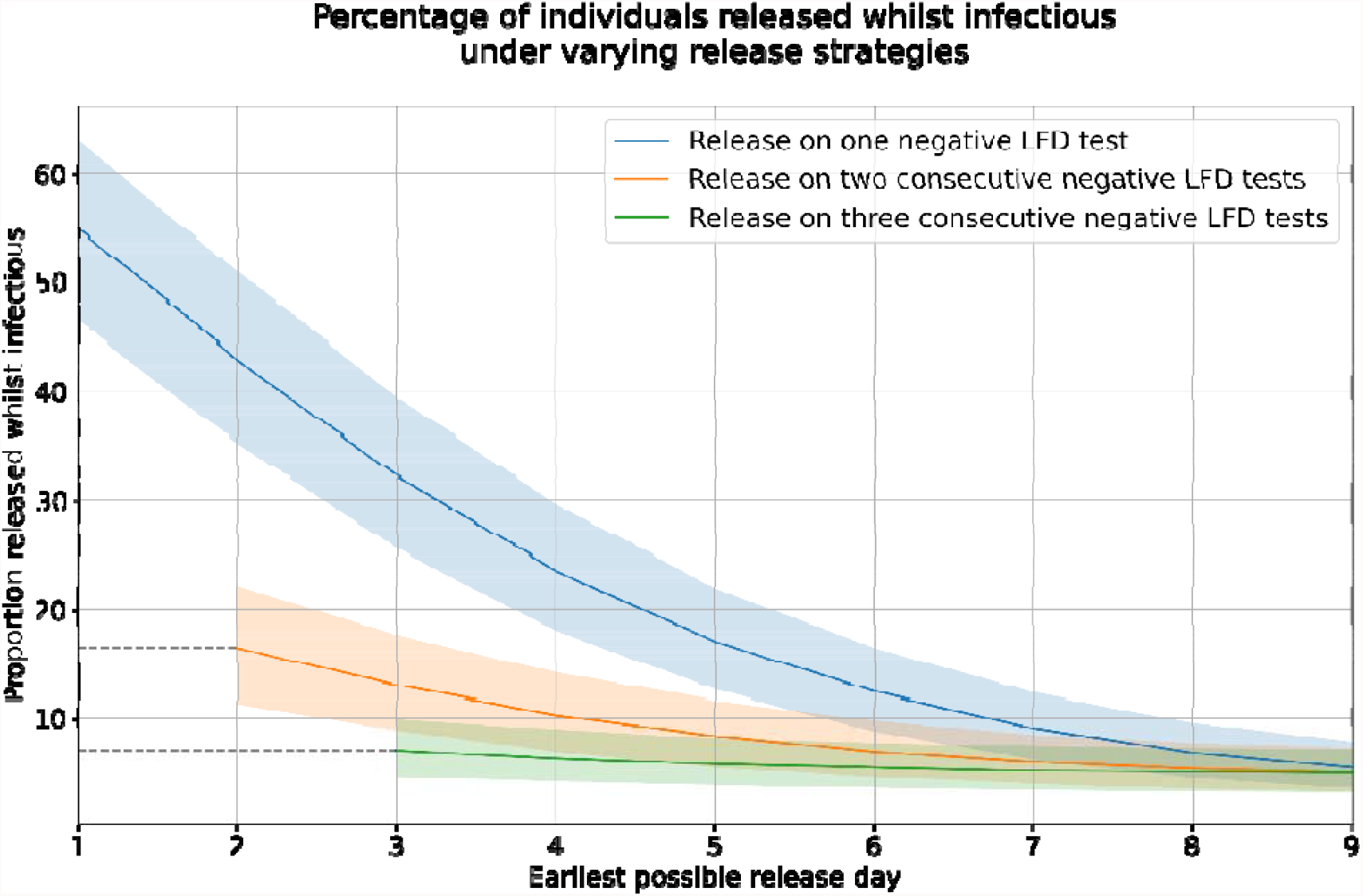
Plot of rates at which model recorded infectious individuals recorded as being released early from each of the considered scenarios (with 95% C.I.). Note dashed lines to signal period in which individuals are ineligible for early release; at least two and three days required where two and three negative tests are required, each 24 hours apart.

**Figure 2:**
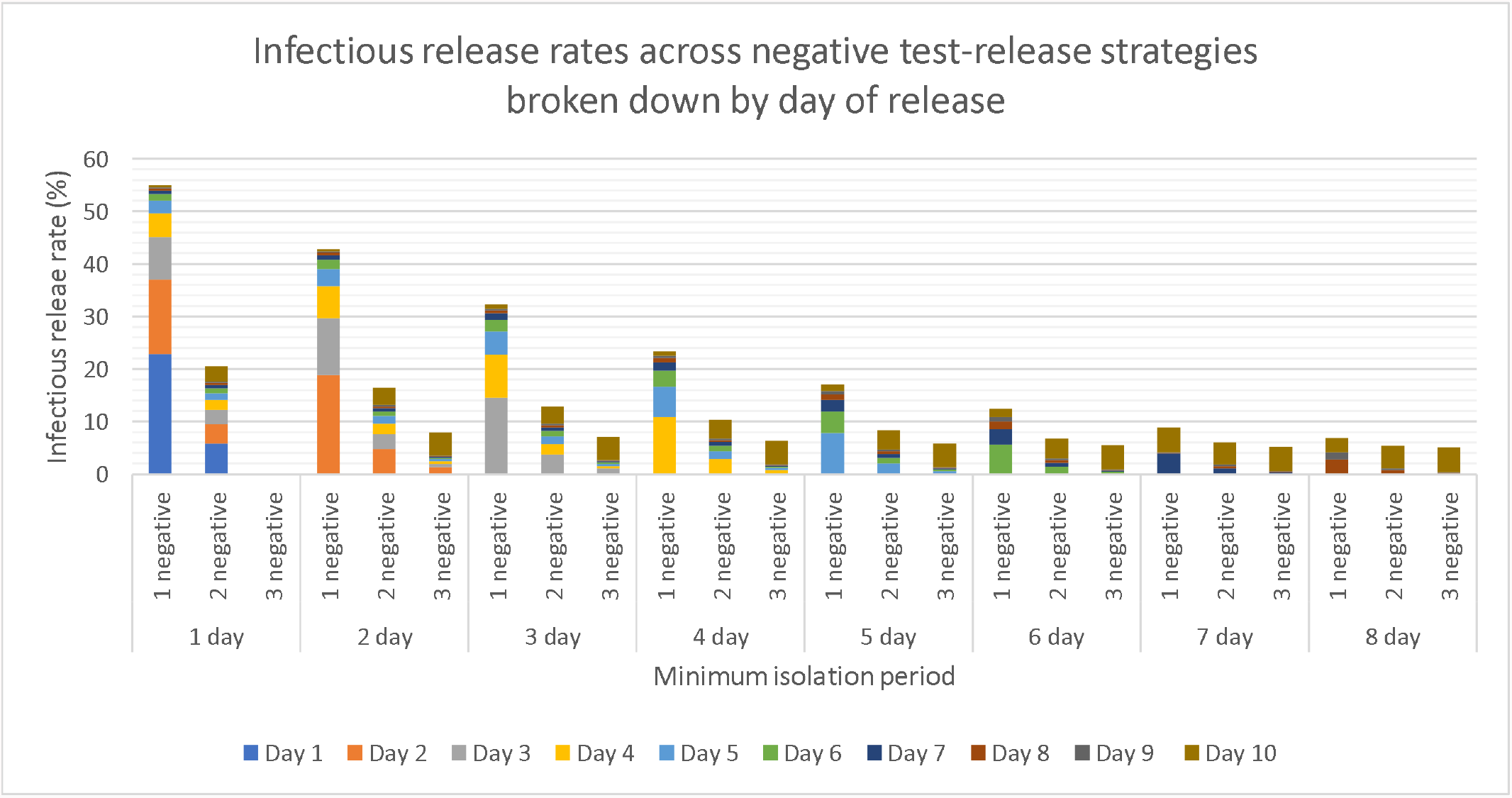
Plots of average release rates at which model reported infectious individuals being released early for each considered scenarios, broken down by day on which infectious individuals are released.

Next, we consider the effect upon time spent infectious following release and excess time spent in isolation. We again see a sizeable decline in time spent infectious following release when LFD testing is being used in conjunction with isolation. This therefore translated into a decline in the potential that isolated individuals may go on to cause secondary cases following release. For 5-day fixed isolation, we see a 47.1% decrease against the scenario where individuals undergo 5 days of isolation and must produce one negative LFD result for early release – bumping up to a decrease of 75.0% and 82.8% when two and three LFD negatives are required respectively. Similar to the above, the fact that we require additional actions to be met to allow release would be expected to present an effect, but the degree of decline is notable. The main trade off for these benefits lay in the additional amount of excess time individuals may spend in isolation following true recovery. Comparing the same instances just described, we would expect to see a 23.8% increase in the average time spent in isolation following recovery when a single negative LFD result is required, increasing to 27.8% and 50.5% when two and three negatives are required respectively.

All the above described effects also interact and should be consider jointly; for instance, if we compare a fixed 10-day isolation period to release from day 8 on condition of two or three sequential LFD negatives, we see that these have nearly identical risks of infectious release. As such, were we looking to stick around a 5% risk of infectious release, we could take this latter approach which would not only allow some individuals to be released one or two days early, but also yield a 29.6% and 27.9% decrease in expected amount of excess time spent in isolation when requiring two and three sequential negatives respectively.

Lastly, we review the potential expense in terms of the number of LFD tests required. As evident from our results, while additional negative results decrease risk of infectious release, figure 3 highlights that this comes at the cost of a considerable number of tests – especially when testing is permitted to start earlier. As such, these scenarios may not be viable where budgets are strict or where there is difficulty in procuring LFD tests. Although, figure 3 also demonstrates how as testing starts later, we would expect average number of tests used to tend towards the minimum of one, two or three depending on the number of negatives results required for early release.

**Figure 3:**
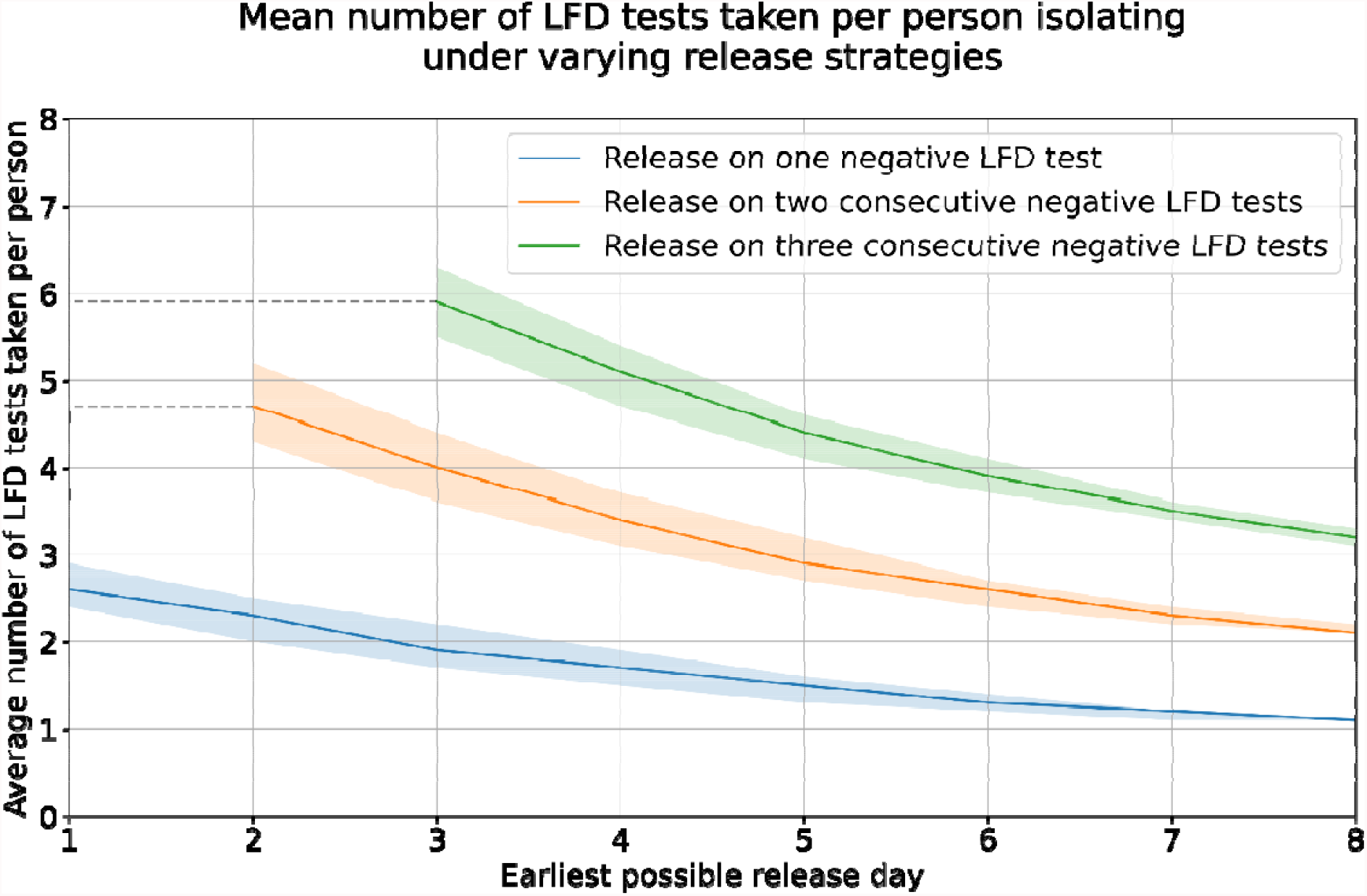
Plots of average number of tests used by simulated individuals in each of the considered scenarios (with 95% C.I.). Note dashed lines to signal period in which individuals are ineligible for early release; at least two and three days required where two and three negative tests are required, each 24 hours apart

While our modelling indicates that the benefit of using LFD testing in conjunction with isolation are sizeable, the choosing between scenarios requires the weighing of the reduction in risk of infectious individuals being released back into the population, against the additional time that recovered individuals may have to spend isolating. Additionally, consideration may need to be paid to the economic cost of provisioning the expected number of LFD tests and whether each scenario may be viable to implement at a national level. In lieu of any LFD testing capability, table 1 shows that a good level of protection may be provided through an extended fixed period of isolation. However, one would have to disregard consideration for excess time spent in isolation that would be expected to be incurred for each individual, being substantial for extended self-isolation periods.

**Table 1:**
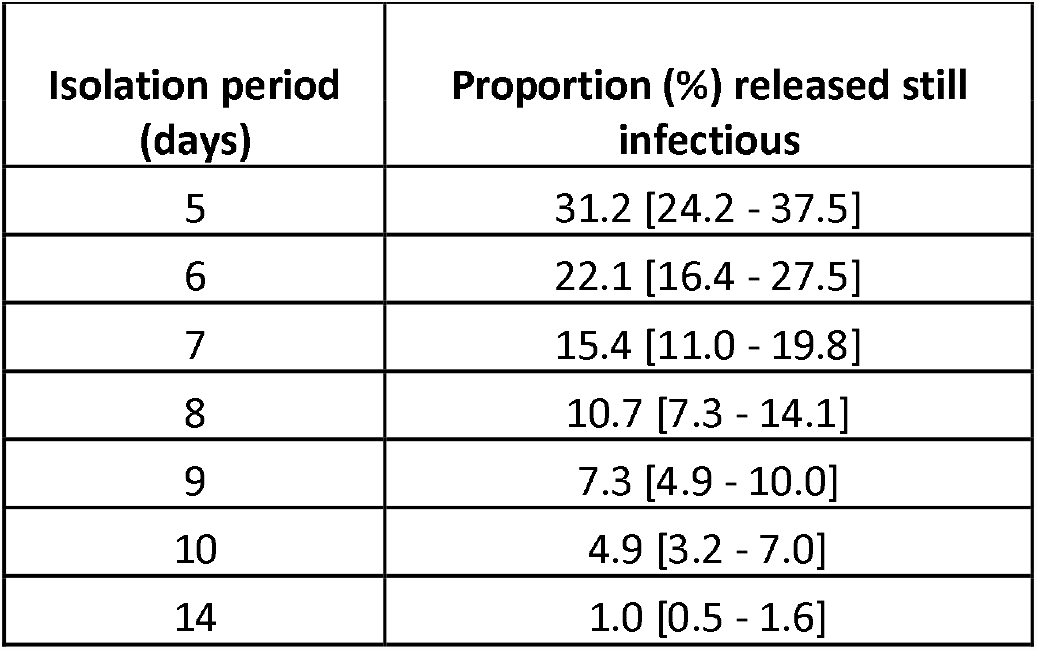
rates at which our model reports infectious individuals would be released were cases required to isolate for a fixed period, absent of any follow-up LFD testing (with 95% C.I.)

| Isolation period (days) | Proportion (%) released still infectious |
| --- | --- |
| 5 | 31.2 [24.2 - 37.5] |
| 6 | 22.1 [16.4 - 27.5] |
| 7 | 15.4 [11.0 - 19.8] |
| 8 | 10.7 [7.3 - 14.1] |
| 9 | 7.3 [4.9 - 10.0] |
| 10 | 4.9 [3.2 - 7.0] |
| 14 | 1.0 [0.5 - 1.6] |

**Table 2:** Time spent infectious outside of isolation and excess time spent in isolation for each individual required to isolate for a fixed period, absent of any follow-up LFD testing (with 95% C.I.)

| Isolation period (days) | Average time spent infectious after early release (hours) | Average excess time spent in isolation (hours) |
| --- | --- | --- |
| 5 | 20.4 [14.4 - 26.7] | 40.0 [34.0 - 47.7] |
| 6 | 14.1 [9.6 - 19.1] | 57.6 [50.2 - 66.9] |
| 7 | 9.6 [6.3 - 13.5] | 77.2 [68.6 - 87.6] |
| 8 | 6.5 [4.2 - 9.4] | 98.1 [88.6 - 109.5] |
| 9 | 4.4 [2.7 - 6.6] | 119.9 [109.7 - 132.1] |
| 10 | 2.9 [1.7 - 4.5] | 142.5 [131.7 - 155.1] |
| 14 | 0.6 [0.3 - 1.0] | 236.1 [224.3 - 249.7] |

**Table 3:** Rates at which our model recorded infectious individuals were being released early from each of the considered scenarios (with 95% C.I.). Note, “N/A” value due to individuals being ineligible for release on day 1 and days 1 and 2 where two and three negative tests are required, each 24 hours apart.

|  | proportion (%) released still infectious |  |  |
| --- | --- | --- | --- |
| Minimum isolation period (days) | One negative test | Two sequential negative tests | Three sequential negative tests |
| 8 | 6.7 [4.2 - 9.8] | 5.3 [3.2 - 8.0] | 5.1 [3.3 - 7.2] |
| 7 | 8.8 [5.9 - 12.3] | 5.9 [3.6 - 8.7] | 5.2 [3.3 - 7.4] |
| 6 | 12.0 [8.5 - 16.1] | 6.8 [4.4 - 9.6] | 5.4 [3.5 - 7.7] |
| 5 | 16.7 [12.3 - 21.6] | 8.1 [5.6 - 11.2] | 5.8 [3.8 - 8.2] |
| 4 | 23.1 [17.7 - 29.1] | 10.0 [7.1 - 13.8] | 6.3 [4.1 - 8.9] |
| 3 | 31.6 [25.7 - 38.9] | 12.6 [9.3 - 17.5] | 7.0 [4.6 - 9.8] |
| 2 | 42.3 [36.2 - 50.7] | 15.9 [11.8 - 22.2] | N/A |
| 1 | 54.3 [47.8 - 63.1] | N/A | N/A |

**Table 4:** Time spent infectious outside of isolation under each of the considered scenarios (with 95% C.I.). Note, “N/A” value due to individuals being ineligible for release on day 1 and days 1 and 2 where two and three negative tests are required, each 24 hours apart.

| Minimum isolation period (days) | Average time spent infectious after early release (hours) |  |  |
| --- | --- | --- | --- |
|  | 1 negative test | 2 sequential negative tests | 3 sequential negative tests |
| 8 | 4.1 [2.5 – 6.2] | 3.2 [2.0 – 5.0] | 3.0 [1.8 – 4.6] |
| 7 | 5.5 [3.4 – 8.1] | 3.6 [2.2 – 5.5] | 3.1 [1.9 – 4.8] |
| 6 | 7.6 [5.0 – 11.0] | 4.2 [2.6 – 6.3] | 3.3 [1.9 – 5.0] |
| 5 | 10.8 [7.2 – 15.2] | 5.1 [3.1 – 7.6] | 3.5 [2.1 – 5.3] |
| 4 | 15.5 [10.7 – 21.1] | 6.5 [3.9 – 9.6] | 3.9 [2.3 – 5.8] |
| 3 | 21.9 [15.6 – 29.0] | 8.4 [5.2 – 12.5] | 4.4 [2.6 – 6.6] |
| 2 | 30.8 [22.8 – 39.7] | 11.2 [7.0 – 16.3] | N/A |
| 1 | 42.4 [32.5 – 53.3] | N/A | N/A |

**Table 5:**
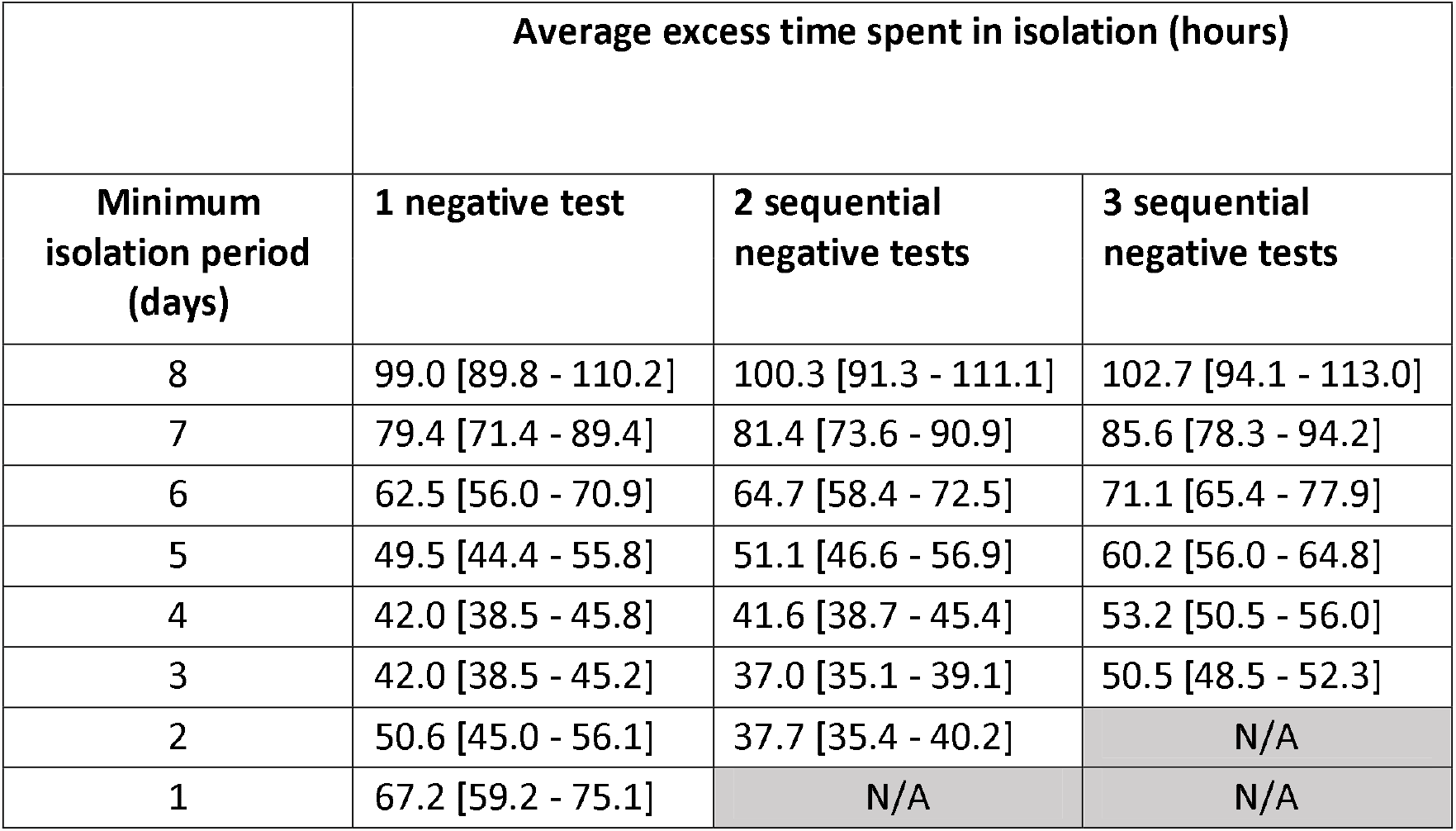
Excess time spent in isolation following while no longer infectious under each of the considered scenarios (with 95% C.I.). Note, “N/A” value due to individuals being ineligible for release on day 1 and days 1 and 2 where two and three negative tests are required, each 24 hours apart.

|  | Average excess time spent in isolation (hours) |  |  |
| --- | --- | --- | --- |
| Minimum isolation period (days) | 1 negative test | 2 sequential negative tests | 3 sequential negative tests |
| 8 | 99.0 [89.8 - 110.2] | 100.3 [91.3 - 111.1] | 102.7 [94.1 - 113.0] |
| 7 | 79.4 [71.4 - 89.4] | 81.4 [73.6 - 90.9] | 85.6 [78.3 - 94.2] |
| 6 | 62.5 [56.0 - 70.9] | 64.7 [58.4 - 72.5] | 71.1 [65.4 - 77.9] |
| 5 | 49.5 [44.4 - 55.8] | 51.1 [46.6 - 56.9] | 60.2 [56.0 - 64.8] |
| 4 | 42.0 [38.5 - 45.8] | 41.6 [38.7 - 45.4] | 53.2 [50.5 - 56.0] |
| 3 | 42.0 [38.5 - 45.2] | 37.0 [35.1 - 39.1] | 50.5 [48.5 - 52.3] |
| 2 | 50.6 [45.0 - 56.1] | 37.7 [35.4 - 40.2] | N/A |
| 1 | 67.2 [59.2 - 75.1] | N/A | N/A |

**Table 6:** Average number of tests used by simulated individuals in each of the considered scenarios (with 95% C.I.). Note, “N/A” value due to individuals being ineligible for release on day 1 and days 1 and 2 where two and three negative tests are required, each 24 hours apart.

|  | Average number of tests used by individuals in isolation |  |  |
| --- | --- | --- | --- |
| Minimum isolation period (days) | 1 negative test | 2 sequential negative tests | 3 sequential negative tests |
| 8 | 1.1 [1.1 - 1.1] | 2.1 [2.1 - 2.2] | 3.2 [3.1 - 3.3] |
| 7 | 1.2 [1.1 - 1.2] | 2.3 [2.2 - 2.4] | 3.5 [3.4 - 3.6] |
| 6 | 1.3 [1.2 - 1.4] | 2.6 [2.4 - 2.7] | 3.9 [3.7 - 4.1] |
| 5 | 1.5 [1.3 - 1.6] | 2.9 [2.7 - 3.2] | 4.4 [4.1 - 4.6] |
| 4 | 1.7 [1.5 - 1.9] | 3.4 [3.1 - 3.7] | 5.1 [4.7 - 5.4] |
| 3 | 1.9 [1.7 - 2.2] | 4.0 [3.6 - 4.4] | 5.9 [5.5 - 6.3] |
| 2 | 2.3 [2.0 - 2.5] | 4.7 [4.3 - 5.2] | N/A |
| 1 | 2.6 [2.4 - 2.9] | N/A | N/A |

## Conclusion

In this work, we have used modelling to illustrate how LFD testing can be implemented to deliver personalised periods of self-isolation which, as opposed to fixed periods of self-isolation, allow us to keep individuals isolated only while they are infectious and thus a public health risk. Our modelling suggests that this approach could allow for a sizeable reduction in the amount of excess time individuals might spend in isolation following recovery (while no longer infectious) compared to longer fixed isolation periods, in addition to a decrease in the average amount of time spent infectious for those released from isolation prior to recovery – hence also the risk of onward transmission. Finally, we see that the use of LFD testing alongside isolation can deliver a reduction in the risk of releasing individuals that are still infectious, while simultaneously decreasing the average time spent in isolation. This effect becomes more prominent as the number of LFD negatives we require to permit early release increases, but such action does also cause additional time to be spent in isolation following recovery and could require significantly more tests to be provisioned for use at a population level. For this reason, we believe the use of two negative LFD tests to provide a suitable compromise between protective effect and cost (both in excess time spent in isolation and financial).

In the absence of available tests, parties should revert to an extended fixed isolation period, the length of which should be set dependent on risk appetite (results herein suggest 5% of infected individuals would be released following a fixed 10-day isolation period, compared to 1% following a fixed 14-day isolation period). In all cases, we urge caution as there is still a chance of residual infectiousness. Should a person be still positive via LFD testing at, for example, day 10 then we would encourage further investigation to confirm infectious status.

## Data Availability

All data has been made available in supplementary materials

