## Supplementary material for "Mitigating isolation: further comparing the effect of LFD testing for early release from self-isolation for COVID-19 cases": S1 - Text

Mitigating isolation: Supplementary material

The table below contains the total output of our model across all considered scenarios.

| Policy | Released infectious (%) | Mean time a released person is infectious for (hours) | Mean released infectious hours per person (hours) | Average excess isolation hours per person released (hours) | Mean excess isolation per person (hours) | Average number of tests per person |
| --- | --- | --- | --- | --- | --- | --- |
| 5-day isolation only | 31.2 [24.2 - 37.5] | 65.0 [58.6 - 71.8] | 20.4 [14.4 - 26.7] | 58.0 [54.3 - 62.6] | 40.0 [34.0 - 47.7] | N/A |
| 6-day isolation only | 22.1 [16.4 - 27.5] | 63.1 [57.2 - 69.4] | 14.1 [9.6 - 19.1] | 73.9 [69.2 - 79.8] | 57.6 [50.2 - 66.9] | N/A |
| 7-day isolation only | 15.4 [11.0 - 19.8] | 61.7 [56.1 - 67.6] | 9.6 [6.3 - 13.5] | 91.2 [85.5 - 98.4] | 77.2 [68.6 - 87.6] | N/A |
| 8-day isolation only | 10.7 [7.3 - 14.1] | 60.5 [55.2 - 66.2] | 6.5 [4.2 - 9.4] | 109.7 [103.1 - 118.1] | 98.1 [88.6 - 109.5] | N/A |
| 9-day isolation only | 7.3 [4.9 - 10.0] | 59.6 [54.4 - 65.0] | 4.4 [2.7 - 6.6] | 129.3 [121.8 - 138.7] | 119.9 [109.7 - 132.1] | N/A |
| 10-day isolation only | 4.9 [3.2 - 7.0] | 58.8 [53.7 - 63.9] | 2.9 [1.7 - 4.5] | 149.8 [141.5 - 160.1] | 142.5 [131.7 - 155.1] | N/A |
| 14-day isolation only | 1.0 [0.5 - 1.6] | 56.6 [51.3 - 61.5] | 0.6 [0.3 - 1.0] | 238.4 [227.8 - 251.1] | 236.1 [224.3 - 249.7] | N/A |
| 10-day isolation, or 1 negative LFD test from day 9 | 5.5 [3.6 - 7.8] | 59.0 [53.8 - 64.4] | 3.3 [2.0 - 5.1] | 126.4 [118.3 - 136.5] | 120.2 [110.1 - 132.3] | 1.0 [1.0 - 1.0] |
| 10-day isolation, or 1 negative LFD test from day 8 | 6.8 [4.5 - 9.5] | 59.6 [54.4 - 64.9] | 4.1 [2.5 - 6.2] | 104.1 [96.4 - 113.7] | 99.0 [89.8 - 110.2] | 1.1 [1.1 - 1.1] |
| 10-day isolation, or 1 negative LFD test from day 7 | 9.0 [6.1 - 12.3] | 60.5 [54.9 - 66.1] | 5.5 [3.4 - 8.1] | 83.5 [76.7 - 92.3] | 79.4 [71.4 - 89.4] | 1.2 [1.1 - 1.2] |
| 10-day isolation, or 1 negative LFD test from day 6 | 12.3 [8.7 - 16.3] | 61.7 [56.0 - 67.6] | 7.6 [5.0 - 11.0] | 65.7 [60.1 - 73.1] | 62.5 [56.0 - 70.9] | 1.3 [1.2 - 1.4] |
| 10-day isolation, or 1 negative LFD test from day 5 | 17.0 [12.5 - 22.0] | 63.2 [57.4 - 69.6] | 10.8 [7.2 - 15.2] | 52.0 [47.8 - 57.4] | 49.5 [44.4 - 55.8] | 1.5 [1.3 - 1.6] |
| 10-day isolation, or 1 negative LFD test from day 4 | 23.5 [18.0 - 29.7] | 65.2 [58.9 - 72.1] | 15.5 [10.7 - 21.1] | 44.2 [41.3 - 47.4] | 42.0 [38.5 - 45.8] | 1.7 [1.5 - 1.9] |
| 10-day isolation, or 1 negative LFD test from day 3 | 32.2 [25.5 - 39.4] | 67.8 [61.4 - 75.2] | 21.9 [15.6 - 29.0] | 44.2 [40.4 - 47.8] | 42.0 [38.5 - 45.2] | 1.9 [1.7 - 2.2] |
| 10-day isolation, or 1 negative LFD test from day 2 | 42.9 [35.2 - 50.9] | 71.4 [64.3 - 79.6] | 30.8 [22.8 - 39.7] | 53.3 [47.1 - 59.6] | 50.6 [45.0 - 56.1] | 2.3 [2.0 - 2.5] |
| 10-day isolation, or 1 negative LFD test from day 1 | 54.9 [46.4 - 63.1] | 76.9 [68.4 - 85.8] | 42.4 [32.5 - 53.3] | 70.7 [62.2 - 78.7] | 67.2 [59.2 - 75.1] | 2.6 [2.4 - 2.9] |
| 10-day isolation, or 2 negative LFD tests from day 8 | 5.1 [3.3 - 7.2] | 58.8 [53.8 - 64.0] | 3.0 [1.8 - 4.7] | 127.1 [119.0 - 137.0] | 120.8 [110.8 - 132.7] | 2.0 [2.0 - 2.0] |
| 10-day isolation, or 2 negative LFD tests from day 7 | 5.4 [3.5 - 7.7] | 59.0 [54.0 - 64.3] | 3.2 [2.0 - 5.0] | 105.5 [98.0 - 114.7] | 100.3 [91.3 - 111.1] | 2.1 [2.1 - 2.2] |
| 10-day isolation, or 2 negative LFD tests from day 6 | 6.0 [3.9 - 8.4] | 59.4 [54.2 - 64.8] | 3.6 [2.2 - 5.5] | 85.5 [79.0 - 93.8] | 81.4 [73.6 - 90.9] | 2.3 [2.2 - 2.4] |
| 10-day isolation, or 2 negative LFD tests from day 5 | 6.9 [4.6 - 9.6] | 60.2 [54.8 - 65.6] | 4.2 [2.6 - 6.3] | 68.0 [62.7 - 74.8] | 64.7 [58.4 - 72.5] | 2.6 [2.4 - 2.7] |
| 10-day isolation, or 2 negative LFD tests from day 4 | 8.3 [5.5 - 11.4] | 61.2 [55.7 - 67.0] | 5.1 [3.1 - 7.6] | 53.7 [50.1 - 58.7] | 51.1 [46.6 - 56.9] | 2.9 [2.7 - 3.2] |
| 10-day isolation, or 2 negative LFD tests from day 3 | 10.2 [6.9 - 14.1] | 62.7 [57.1 - 68.9] | 6.5 [3.9 - 9.6] | 43.7 [41.5 - 46.7] | 41.6 [38.7 - 45.4] | 3.4 [3.1 - 3.7] |
| 10-day isolation, or 2 negative LFD tests from day 2 | 12.9 [8.8 - 17.6] | 64.9 [59.2 - 71.7] | 8.4 [5.2 - 12.5] | 39.0 [37.6 - 40.3] | 37.0 [35.1 - 39.1] | 4.0 [3.6 - 4.4] |
| 10-day isolation, or 2 negative LFD tests from day 1 | 16.3 [11.2 - 22.0] | 67.8 [61.9 - 74.9] | 11.2 [7.0 - 16.3] | 39.7 [37.5 - 42.3] | 37.7 [35.4 - 40.2] | 4.7 [4.3 - 5.2] |
| 10-day isolation, or 2 negative LFD tests from day 0 | 20.4 [14.1 - 27.1] | 72.2 [65.5 - 79.6] | 14.9 [9.4 - 21.4] | 44.4 [40.6 - 48.7] | 42.2 [38.5 - 46.3] | 5.5 [5.1 - 6.0] |
| 10-day isolation, or 3 negative LFD tests from day 7 | 5.0 [3.2 - 7.1] | 58.8 [53.7 - 63.9] | 3.0 [1.8 - 4.6] | 128.1 [120.3 - 137.7] | 121.8 [112.0 - 133.5] | 3.0 [3.0 - 3.0] |
| 10-day isolation, or 3 negative LFD tests from day 6 | 5.1 [3.3 - 7.2] | 58.8 [53.7 - 64.0] | 3.0 [1.8 - 4.6] | 108.0 [101.0 - 116.6] | 102.7 [94.1 - 113.0] | 3.2 [3.1 - 3.3] |
| 10-day isolation, or 3 negative LFD tests from day 5 | 5.2 [3.3 - 7.4] | 59.0 [54.0 - 64.1] | 3.1 [1.9 - 4.8] | 90.0 [84.1 - 97.2] | 85.6 [78.3 - 94.2] | 3.5 [3.4 - 3.6] |
| 10-day isolation, or 3 negative LFD tests from day 4 | 5.4 [3.5 - 7.7] | 59.2 [54.0 - 64.6] | 3.3 [1.9 - 5.0] | 74.8 [70.2 - 80.4] | 71.1 [65.4 - 77.9] | 3.9 [3.7 - 4.1] |
| 10-day isolation, or 3 negative LFD tests from day 3 | 5.8 [3.8 - 8.2] | 59.6 [54.5 - 64.9] | 3.5 [2.1 - 5.3] | 63.3 [60.2 - 66.8] | 60.2 [56.0 - 64.8] | 4.4 [4.1 - 4.6] |
| 10-day isolation, or 3 negative LFD tests from day 2 | 6.3 [4.1 - 8.9] | 60.4 [55.2 - 66.1] | 3.9 [2.3 - 5.8] | 56.0 [54.2 - 57.8] | 53.2 [50.5 - 56.0] | 5.1 [4.7 - 5.4] |
| 10-day isolation, or 3 negative LFD tests from day 1 | 7.0 [4.6 - 9.8] | 61.7 [56.1 - 67.6] | 4.4 [2.6 - 6.6] | 53.1 [52.0 - 54.2] | 50.5 [48.5 - 52.3] | 5.9 [5.5 - 6.3] |
| 10-day isolation, or 3 negative LFD tests from day 0 | 8.0 [5.2 - 11.1] | 63.5 [58.1 - 69.8] | 5.1 [3.0 - 7.7] | 53.1 [52.4 - 53.9] | 50.4 [48.8 - 52.0] | 6.8 [6.4 - 7.2] |

The next table contains the infectious release rates for each of the above scenarios, broken down into expected proportion to be released on each of the viable days:

|  |  |  | Average day released on (%) | | | | | | | | | |
| --- | --- | --- | --- | --- | --- | --- | --- | --- | --- | --- | --- | --- |
|  |  |  | Day 1 | Day 2 | Day 3 | Day 4 | Day 5 | Day 6 | Day 7 | Day 8 | Day 9 | Day 10 |
| Minimum isolation period | 1 day | 1 negative | 22.9 | 14.1 | 8.1 | 4.5 | 2.4 | 1.3 | 0.7 | 0.4 | 0.2 | 0.4 |
|  |  | 2 negatives | 5.8 | 3.6 | 2.8 | 1.9 | 1.3 | 0.9 | 0.6 | 0.4 | 0.2 | 3 |
|  |  | 3 negatives | NA | NA | NA | NA | NA | NA | NA | NA | NA | NA |
|  | 2 day | 1 negative | NA | 18.8 | 10.9 | 6 | 3.3 | 1.7 | 0.9 | 0.5 | 0.2 | 0.5 |
|  |  | 2 negatives | NA | 4.8 | 2.8 | 2 | 1.4 | 0.9 | 0.6 | 0.4 | 0.3 | 3.2 |
|  |  | 3 negatives | NA | 1.2 | 0.7 | 0.5 | 0.4 | 0.3 | 0.2 | 0.1 | 0.1 | 4.4 |
|  | 3 day | 1 negative | NA | NA | 14.6 | 8.1 | 4.4 | 2.3 | 1.2 | 0.6 | 0.3 | 0.7 |
|  |  | 2 negatives | NA | NA | 3.7 | 2 | 1.5 | 1 | 0.6 | 0.4 | 0.3 | 3.3 |
|  |  | 3 negatives | NA | NA | 1 | 0.5 | 0.4 | 0.3 | 0.2 | 0.1 | 0.1 | 4.5 |
|  | 4 day | 1 negative | NA | NA | NA | 10.8 | 5.8 | 3.1 | 1.6 | 0.8 | 0.4 | 0.9 |
|  |  | 2 negatives | NA | NA | NA | 2.8 | 1.5 | 1.1 | 0.7 | 0.4 | 0.3 | 3.5 |
|  |  | 3 negatives | NA | NA | NA | 0.7 | 0.4 | 0.3 | 0.2 | 0.1 | 0.1 | 4.6 |
|  | 5 day | 1 negative | NA | NA | NA | NA | 7.8 | 4.1 | 2.2 | 1.1 | 0.6 | 1.2 |
|  |  | 2 negatives | NA | NA | NA | NA | 2 | 1.1 | 0.7 | 0.5 | 0.3 | 3.7 |
|  |  | 3 negatives | NA | NA | NA | NA | 0.5 | 0.3 | 0.2 | 0.1 | 0.1 | 4.6 |
|  | 6 day | 1 negative | NA | NA | NA | NA | NA | 5.6 | 2.9 | 1.5 | 0.8 | 1.6 |
|  |  | 2 negatives | NA | NA | NA | NA | NA | 1.4 | 0.7 | 0.5 | 0.3 | 3.9 |
|  |  | 3 negatives | NA | NA | NA | NA | NA | 0.4 | 0.2 | 0.1 | 0.1 | 4.7 |
|  | 7 day | 1 negative | NA | NA | NA | NA | NA | NA | 3.9 | 0.1 | 0.1 | 4.7 |
|  |  | 2 negatives | NA | NA | NA | NA | NA | NA | 1 | 0.5 | 0.3 | 4.2 |
|  |  | 3 negatives | NA | NA | NA | NA | NA | NA | 0.3 | 0.1 | 0.1 | 4.7 |
|  | 8 day | 1 negative | NA | NA | NA | NA | NA | NA | NA | 2.7 | 1.4 | 2.8 |
|  |  | 2 negatives | NA | NA | NA | NA | NA | NA | NA | 0.7 | 0.3 | 4.4 |
|  |  | 3 negatives | NA | NA | NA | NA | NA | NA | NA | 0.2 | 0.1 | 4.8 |
